# Seropositivity in blood donors and pregnant women during the first year of SARS-CoV-2 transmission in Stockholm, Sweden

**DOI:** 10.1101/2020.12.24.20248821

**Authors:** Xaquin Castro Dopico, Sandra Muschiol, Murray Christian, Leo Hanke, Daniel J. Sheward, Nastasiya F. Grinberg, Joanna Rorbach, Gordana Bogdanovic, Gerald M. Mcinerney, Tobias Allander, Chris Wallace, Ben Murrell, Jan Albert, Gunilla B. Karlsson Hedestam

## Abstract

In Sweden, social restrictions to contain SARS-CoV-2 have to date primarily relied upon voluntary adherence to a set of recommendations and strict lockdowns/regulations have not been enforced, potentially affecting viral dissemination. To understand the levels of past SARS-CoV-2 infection in the Stockholm population before the start of mass vaccinations, healthy blood donors and pregnant women (*n=*5,100) were sampled at random between 14^th^ March 2020-28^th^ February 2021. All individuals (*n=*200/sampling week) were screened for anti-SARS-CoV-2 spike (S) trimer- and RBD-specific IgG responses and the results were compared with those from historical controls (*n=*595). Data were modelled using a probabilistic Bayesian framework that considered individual responses to both viral antigens. We found that after a steep rise at the start of the pandemic, the seroprevalence trajectory increased more steadily (over summer) in approach to the winter second-wave of infections, approaching 15% of all adults surveyed by mid-December 2020. The population seropositivity rate again increased more rapidly as cases rose over the winter period. By the end of February 2021, ∼19% (∼one-in-five) in this study group tested seropositive. Notably, 96% of random seropositive samples screened (*n*=56), displayed virus neutralizing responses, with titers comparable to those engendered by recently approved mRNA vaccines, supporting that milder infections generally provoke a competent B cell response. These data offer baseline information about the level of seropositivity in this group of active adults in the Stockholm metropolitan area following a full year of SARS-CoV-2 transmission and prior to the introduction of vaccines.

**Structured abstract:** *Objectives:* Sweden did not enforce social lockdown in response to the SARS-CoV-2 pandemic. Therefore, we sought to determine the proportion of seropositive healthy, active adults in Stockholm, the country’s most populous region. Random sampling (of blood donors and pregnant women) was carried out during the first year following virus emergence in the country and prior to vaccination of the general adult population – allowing for an estimate of seroprevalence in response to natural infection.

*Design:* In this cross-sectional prospective study, otherwise-healthy blood donors (*n=*2,600) and pregnant women(*n=*2,500) were sampled at random for consecutive weeks (at four intervals) between 14^th^ March and 28^th^ February 2021. Sera from all participants and a cohort of historical controls (*n=*595) were screened for IgG responses against trimers of the SARS-CoV-2 spike (S) glycoprotein and the smaller receptor-binding domain (RBD). As a complement to standard analytical approaches, a probabilistic (cut-off-independent) Bayesian framework that assigns likelihood of past infection was used to analyze data over time. The study was carried out in accordance with Swedish Ethical Review Authority: registration number 2020-01807.

*Setting:* Healthy participant samples were selected from their respective pools at random through Karolinska University Hospital.

*Participants:* None of the participants were symptomatic at sampling. No additional metadata was available from the samples.

*Results:* Blood donors and pregnant women showed a similar seroprevalence. After a steep rise at the start of the pandemic, the seroprevalence trajectory increased steadily in approach to the winter second-wave of infections, approaching 15% of all individuals surveyed by 13^th^ December 2020. By the end of February 2021, when deaths were in decline and at low levels following their winter peak, 19% of the population tested seropositive. Notably, 96% of seropositive healthy donors screened (*n=*56) developed neutralizing antibody responses at titers comparable to, or higher than those observed in clinical trials of SARS-CoV-2 spike mRNA vaccination, supporting that mild infection engenders a competent B cell response.

*Conclusions:* These data indicate that in the year since the start of community transmission, seropositivity levels in metropolitan Stockholm had reached approximately one-in-five persons, providing important baseline seroprevalence information prior to the start of vaccination.

## Introduction

Densely populated areas – such as the Stockholm region of 2.37 million people (975,000 within city limits) – have facilitated the spread of SARS-CoV-2. Evidence suggests that transmission can be curtailed by imposing restrictions on leisure and business activities, as well as by mask usage and contact tracing^1–4^. In contrast to the majority of comparable countries, Sweden has favoured a strategy in which individuals are encouraged to adhere to a set of basic public health recommendations, while society has remained largely open. Only recently – faced with a winter second wave – did the government opt for earlier closing hours for some businesses. The country has reported a significantly higher burden on the public healthcare system to date than its Scandinavian neighbours (Fig. 1A), and there are many reasons for why this might be the case.

**Figure 1:**
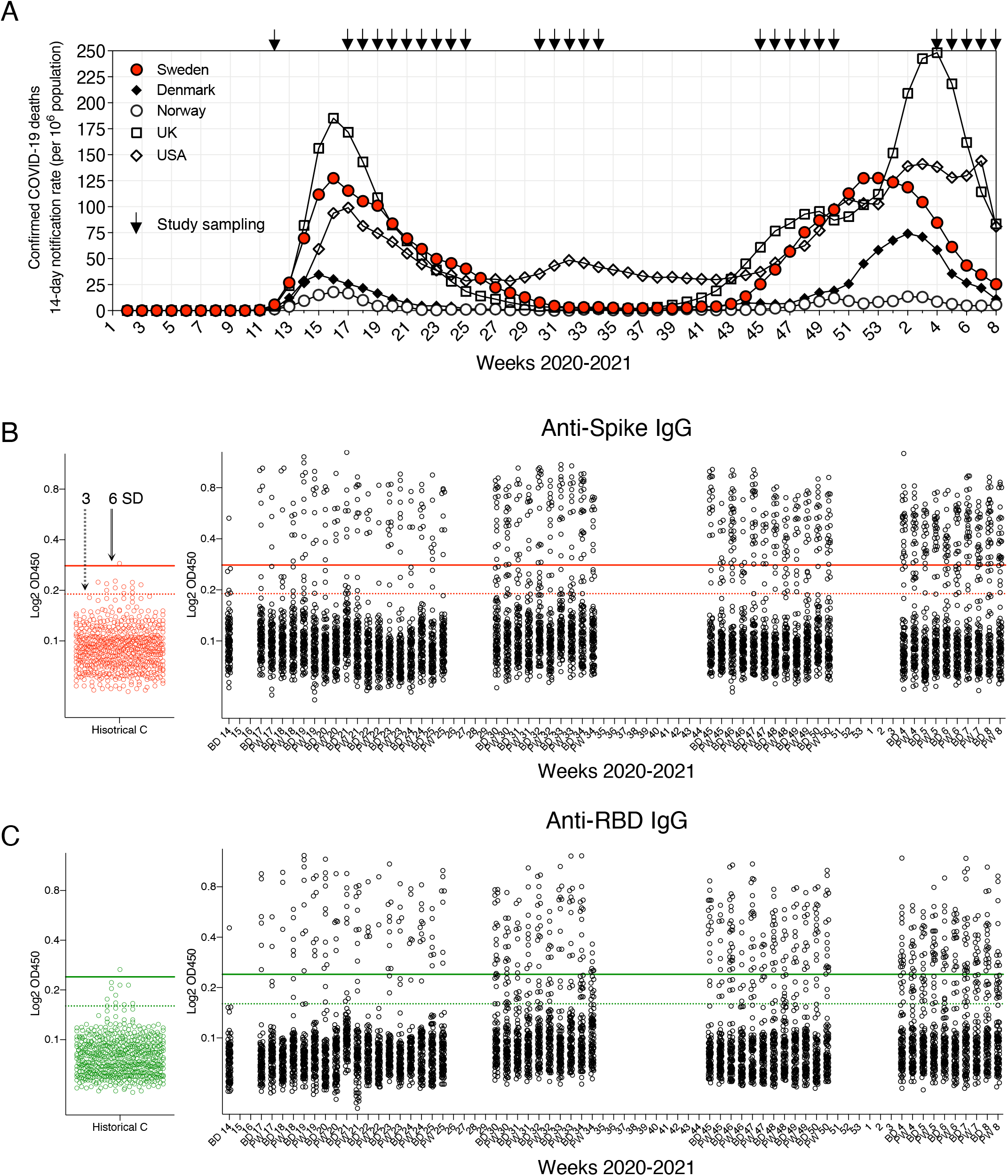
SARS-CoV-2 seropositivity estimates in Stockholm: March 2020-February 2021. **(A)** Population-adjusted COVID-19 deaths for selected countries during the pandemic. **(B)** Anti-S IgG responses in blood donors (BD), pregnant women (PW) and *n=*595 historical control sera. 100 BD and 100 PW samples were analyzed per sampling week alongside negative controls (cut-out plot red points). Conventional 3 and 6 SD (from the mean of negative control values) assay cut-offs are shown as dashed and solid lines, respectively. **(D)** Anti-RBD IgG responses in blood donors (BD), pregnant women (PW) and *n=*595 historical control sera (cut-out plot green points). Conventional 3 and 6 SD assay cut-offs are shown as dashed and solid lines, respectively.

Serological enumeration of past infections is critical for estimating viral spread and understanding characteristics of the adaptive immune response to an emerging pathogen^5^. Antibody testing is also important for optimal planning of vaccination campaigns (especially when doses are limited) and could help demonstrate “immunity” on official documentation. However, as illustrated by the current pandemic, not all antibody tests are of equal sensitivity and specificity (S&S), and closer scrutiny of assays during regulatory approval is needed to improve public health measures and our understanding of Covid-19^6^.

To monitor seropositivity in the Stockholm region, we developed highly S&S SARS-CoV-2 antibody tests based on native-like spike glycoprotein antigens^7,8^ (alongside a diagnostic clinical laboratory responsible for monitoring seropositivity during the pandemic^9^) and novel (cut-off-independent) statistical methods, and applied them to random, healthy adults in the Stockholm region throughout 2020 and early 2021. Such methods, able to survey antibody responses to multiple antigens and improve upon strictly thresholding a continuous variable, are critical for accurate seropositivity estimates at individual and population levels. This is especially important in the case of SARS-CoV-2, as asymptomatic/mild infections generate antibody responses of varying titers that can be difficult to classify^9–13^ and individual titers wane from peak levels over time11,14–16.

We chose to survey anti-S IgG responses as these are the best indicators of past SARS-CoV-2 infection at the population level (i.e. present in >91% of PCR-positive cases)^17,18^. Spike harbours the ACE2 receptor binding domain (RBD) and is the major target of the neutralizing antibody response, central to vaccine efforts. For example, anti-nucleocapsid (N) antibodies were not detectable in a subset of S-seropositive individuals^9,19–22^. Forboth S, N and RBD, the levels of antibodies in circulation were shown to increase with worsening disease severity^10,13,23,24^, as also reported for SARS-CoV and MERS^25,26^.

Blood donors and pregnant women, studied here, are important clinical groups, and pregnant women in particular require further study in relation to infectious disease^27^. Moreover, both groups represent good and accessible proxies for adult population health, being generally working-age, mobile members of society -without being enriched for individuals at especially high-risk of SARS-CoV-2 infection, such as healthcare workers or public transportation employees, where seroprevalence may be higher^28^. Similarly, seroprevalence may be higher in children and lower in the elderly – with complementary studies much needed.

All healthy individuals in this study (*n*=5,100) were over the age of 18 and symptom-free at sample collection. For ethical reasons, their ages and genders are not revealed, nor whether any participant previously tested positive for the virus. Thus, while we here provide an unbiased assessment of past SARS-CoV-2 infection in two important groups in Stockholm before the start of mass adult vaccinations (in response to natural infection), future studies in other cohorts are needed to generate more integrated public health strategies for managing the pandemic.

We follow seropositivity – IgG – in these study groups across the first and second waves of infections, informing the development of population immunity over time in response to natural infection and shortly before Covid-19 vaccines entered the general population.

## Materials and methods

### Human samples and ethical declaration

Anonymized samples (*n=*5,100 total) from blood donors (*n=*100/sampling week) and pregnant women (*n=*100/sampling week) were randomly selected from their respective pools by the department of Clinical Microbiology, Karolinska University Hospital. During 2020, 18,963 pregnant women were under the care of Karolinska University Hospital, averaging 365 per week, of which 100 (*∼*30%) were selected at random from each sampling week. Blood donations totalled 66,596 during 2020, averaging 1,281 per week and here weekly sampling approximated 8% of the pool. No study participant was analysed more than once over the course of the study. Blood donor and pregnant women samples were not collected during weeks 26-29, 35-45, 51-53 2020 and 1-3 2021 for logistical reasons. Samples were collected from 26 weeks out of the 47 weeks the study ran. Blood donors (*n*=595) collected through the same channels one year previously (Spring 2019) were randomly selected for use as assay negative controls.

No metadata, such as age or sex information were available for the samples in the study. Blood donors must be over 50 kg and the age of 18, while no upper age limit is established in Stockholm. Pregnant women were sampled as part of routine screening for infectious diseases during the first trimester of pregnancy. Whether a study participant had previously tested positive for SARS-CoV-2 PCR is unknown. Blood donors are required to be healthy for a minimum of two weeks before a donation and all participants reported symptom-free at sample collection. We cannot exclude that a very small number of blood donor samples from 2021 were from individuals who had received a Covid-19 vaccine. Healthcare workers in the region were receiving first doses at the time of writing, along with elderly persons; however, mass vaccinations of adults (including pregnant women) had not yet begun^29,30^.

The use of study samples was approved by the Swedish Ethical Review Authority (registration no. 2020-01807). Stockholm County death and Swedish mortality data was sourced from the ECDC, EU (https://www.ecdc.europa.eu/en/covid-19/data) and were current at the time of writing.

### Serum sample processing

Blood samples were collected by the attending clinical team and serum isolated by the Department of Clinical Microbiology. Samples were anonymized, barcoded and stored at -20°C until use. Serum samples were not heat-inactivated for ELISA protocols.

### SARS-CoV-2 antigen generation

The plasmid for expression of the SARS-CoV-2 prefusion-stabilized spike ectodomain with a C-terminal T4 fibritin trimerization motif was obtained from Wrapp *et al*.^8^ and produced as in Hanke *et al*^7^. Briefly, the plasmid was used to transiently transfect FreeStyle 293F cells using FreeStyle MAX reagent (Thermo Fisher Scientific). The ectodomain was purified from filtered supernatant on Streptactin XT resin (IBA Lifesciences), followed by size-exclusion chromatography on a Superdex 200 in 5 mM Tris pH 8, 200 mM NaCl. The RBD domain (RVQ – QFG) of SARS-CoV-2 was cloned upstream of a sortase A recognition site (LPETG) and a 6xHIS tag and expressed in 293F cells as described above. RBD-HIS was purified from filtered supernatant on His-Pur Ni-NTA resin (Thermo Fisher Scientific), followed by size-exclusion chromatography on a Superdex 200.

### Anti-SARS-CoV-2 ELISA

96-well ELISA plates (Nunc MaxiSorp) were coated with freshly prepared SARS-CoV-2 S trimers or the RBD (100 µl of 1 ng/µl) in PBS for 15 h at 4°C. Plates were washed six times with PBS-Tween-20 (0.05%) and blocked using PBS-5% no-fat milk (Sigma). Human serum samples were thawed at room temperature, diluted, vortexed and incubated in blocking buffer for 1 h (4°C) before plating to block non-specific binding. Serum samples were incubated for 15 h at 4°C to allow low-affinity binding interactions, before washing as before. Secondary HRP-conjugated anti-human antibodies were diluted in blocking buffer and incubated with samples for 1 hour at 4°C. Plates were washed a final time before development with TMB Stabilized Chromogen kept at 4°C (Invitrogen). The reaction was stopped using 1M sulphuric acid and optical density (OD) values were measured at 450 nm using an Asys Expert 96 ELISA reader (Biochrom Ltd.). Secondary antibodies (from Southern Biotech) and dilutions used: goat anti-human IgG (2014-05) at 1:10,000. All assays were developed for their fixed time and negative control samples were run alongside test samples in all assays. Raw 450nm optical density (OD) data were log transformed for statistical analyses and “no samples control wells” across plates were used for assay-assay normalization. Anti-SARS-CoV-2 S and RBD IgG were detectable at up to 1:20,000 serum dilution using this assay^9^ and all study samples were here run at 1:100 dilution.

### *In vitro* virus neutralization assay

Pseudotyped viruses were generated by the co-transfection of HEK293T cells with plasmids encoding the SARS-CoV-2 spike protein harboring an 18 amino acid truncation of the cytoplasmic tail^8^; a plasmid encoding firefly luciferase; a lentiviral packaging plasmid (Addgene 8455) using Lipofectamine 3000 (Invitrogen). Media was changed 12-16 hours post-transfection and pseudotyped viruses harvested at 48- and 72-hours, filtered through a 0.45 µm filter and stored at -80°C until use. Pseudotyped neutralization assays were adapted from protocols validated to characterize the neutralization of HIV, but with the use of ACE2-expressing HEK293T cells. Briefly, pseudotyped viruses sufficient to generate ∼100,000 RLUs were incubated with serial dilutions of heat-inactivated serum for 60 min at 37°C. Approximately 15,000 HEK293T-ACE2 cells were then added to each well and the plates incubated at 37°C for 48 hours. Luminescence was measured using Bright-Glo (Promega) according to the manufacturer’s instructions on a GM-2000 luminometer (Promega) with an integration time of 0.3s. The neutralization assay limit of detection was at 1:45 serum dilution.

### Probabilistic seroprevalence estimations

Prior to analysis, each sample OD was standardized by dividing by the mean OD of “no sample control” wells on that plate or other plates run on the same day – to reduce batch variation. This resulted in more similar distributions for 2019 blood donor samples with 2020 blood donors and pregnant volunteers. Our Bayesian approach is presented in detail in Christian and Murrell^31^. Briefly, we used a logistic regression over anti-RBD and -S training data (from *n=*595 historical blood donor controls and *n*=138 SARS-CoV-2 PCR+ individuals across the clinical spectrum – all of whom developed anti-S and -RBD IgG^9^) to model the relationship between the ELISA measurements and the probability that a sample is antibody-positive. We adjusted for the training data class proportions and used these adjusted probabilities to inform the seroprevalence estimates for each time point. Given that the population seroprevalence cannot increase dramatically from one week to the next, we constructed a prior over seroprevalence trajectories using a transformed Gaussian Process, and combined this with the individual class-balance adjusted infection probabilities for each donor to infer the posterior distribution over seroprevalence trajectories. We compared our Bayesian approach to the output of more established probabilistic algorithms (specifically, an ensemble learner from support vector machines (SVM) and linear discriminant analysis (LDA)) that we have previously developed for ELISA measurements^9^.

The sensitivity and specificity (S&S) of our IgG ELISA assays were:

Spike 3SD: *100% (95% CI [97*.*5-100*.*0]) & 99*.*0% (95% CI [98*.*6-99*.*0])*.

Spike 6SD: *100% (95% CI [97*.*5-100*.*0]) & 99*.*9% (95% CI [99*.*6-100*.*0])*.

RBD 3SD: *100% (95% CI [97*.*5-100*.*0]) & 99*.*0% (95% CI [98*.*4-99*.*4])*.

RBD 6SD: *98*.*0% (95% CI [94*.*2-99*.*3]) & 99*.*9% (95% CI [99*.*6-100*.*0])*.

SVM-LDA: *99*.*3% (95% CI [96*.*3-100*.*0) & 100*.*0% (95% CI [99*.*8-100*.*0])*.

To compute confidence intervals for S&S, we dichotomized predictions of seropositivity at prob > 0.5 or <= 0.5 and computed average sensitivity, specificity, and 95% confidence intervals for each fold in the cross validation via Wilson’s method before averaging over all folds. The Bayesian model does not make assignments of seropositivity to individual samples, but rather integrates over the uncertainty in relationship between infection and OD450, as well as the uncertainty due to sampling, to provide population-level seroprevalence estimates. Thus, S&S cannot be calculated for this approach.

## Results

Blood donor and pregnant women serum samples (*n=*100 of each per sampling week) were selected at random from their respective pools and the IgG response against SARS-CoV-2 S glycoprotein trimers and the smaller RBD subunit was measured in all sera using established ELISA assays extensively validated using sera from confirmed infections^9^ (Fig. 1B and C). Anti-S and RBD responses are highly correlated, with lower titers generally observed for the smaller RBD^9^. Test samples were run alongside historical (SARS-CoV-2-negative) control sera (*n=*595 blood donors from spring 2019) throughout the study and assay-assay variation was controlled by normalization to signal in “no sample control” wells. Seropositivity according to conventional 3 or 6 standard deviations (SD) (from the mean of negative control sera) assay cut-offs is presented in Table 1 and Fig. 2A). Notably, seropositivity was not significantly different between blood donors and pregnant women throughout the study (Fig. 2 B and C).

**Table 1:** *IgG seropositivity to S and RBD in blood donors and pregnant women* following virus emergence.

| ***Weeks 2020-2021*** | **Bayesian estimate** | **S IgG (3SD)** | **RBD IgG (3SD)** | **S IgG (6SD)** | **RBD IgG (6SD)** |
| --- | --- | --- | --- | --- | --- |
| **2020** |  |  |  |  |  |
| **14: Mar 30<sup>th</sup>-Apr 5<sup>th</sup>** | 2.4 | 2.5 | 0.5 | 0.5 | 0.5 |
|  | *Sampling gap* |  |  |  |  |
| **17: Apr 20-27<sup>th</sup>** | 4.8 | 6.0 | 5.0 | 4.5 | 4.0 |
| **18: Apr 27<sup>th</sup>-May 3<sup>rd</sup>** | 5.4 | 8.0 | 5.0 | 5.0 | 4.0 |
| **19: May 4-10<sup>th</sup>** | 5.9 | 11.5 | 8.0 | 9.0 | 8.0 |
| **20: May 11-17<sup>th</sup>** | 6.3 | 8.0 | 9.0 | 7.0 | 8.0 |
| **21: May 18-24<sup>th</sup>** | 6.7 | 12.0 | 10.0 | 8.0 | 7.5 |
| **22: May 25<sup>th</sup>-31<sup>st</sup>** | 7.0 | 4.5 | 3.5 | 4.0 | 3.5 |
| **23: Jun 1<sup>st</sup>-7<sup>th</sup>** | 7.3 | 10.5 | 9.0 | 9.5 | 7.0 |
| **24: Jun 8-14<sup>th</sup>** | 7.8 | 8.0 | 8.0 | 6.5 | 7.0 |
| **25: Jun 15<sup>th</sup>-21<sup>st</sup>** | 8.3 | 8.5 | 7.0 | 7.5 | 7.0 |
|  | *Sampling gap* |  |  |  |  |
| **30: Jul 20-26<sup>th</sup>** | 11.4 | 22.0 | 20.5 | 17.0 | 14.5 |
| **31: Jul 27<sup>th</sup>-Aug 2<sup>nd</sup>** | 11.9 | 14.5 | 13.5 | 10.5 | 9.0 |
| **32: Aug 3<sup>rd</sup>-9<sup>th</sup>** | 12.3 | 18.0 | 16.0 | 13.5 | 10.5 |
| **33: Aug 10-16<sup>th</sup>** | 12.5 | 15.5 | 12.5 | 12.5 | 10.0 |
| **34: Aug 17<sup>th</sup>-23<sup>rd</sup>** | 12.7 | 20.0 | 20.0 | 16.5 | 9.5 |
|  | *Sampling gap* |  |  |  |  |
| **45: Nov 2<sup>nd</sup>-8<sup>th</sup>** | 14.1 | 15.5 | 14.0 | 12.5 | 11.0 |
| **46: Nov 9-15<sup>th</sup>** | 14.2 | 21.0 | 17.0 | 16.5 | 13.5 |
| **47: Nov 16<sup>th</sup>-22<sup>nd</sup>** | 14.3 | 17.5 | 16.0 | 17.0 | 12.0 |
| **48: Nov 23<sup>rd</sup>-29<sup>th</sup>** | 14.5 | 20.5 | 18.0 | 16.0 | 12.5 |
| **49: Nov 30<sup>th</sup>-Dec 6<sup>th</sup>** | 14.7 | 16.5 | 14.5 | 12.5 | 12.0 |
| **50: Dec 7-13<sup>th</sup>** | 14.8 | 20.0 | 17.5 | 18.0 | 15.5 |
| **2021** | *Sampling gap* |  |  |  |  |
| **4: Jan 25<sup>th</sup>-31<sup>st</sup>** | 18.2 | 28.5 | 22.0 | 23.5 | 15.0 |
| **5: Feb 1<sup>st</sup>-7<sup>th</sup>** | 18.5 | 23.5 | 23.0 | 21.5 | 13.0 |
| **6: Feb 8-14<sup>th</sup>** | 18.8 | 25.5 | 19.5 | 19.0 | 15.5 |
| **7: Feb 15<sup>th</sup>-21<sup>st</sup>** | 19.0 | 28.5 | 25.0 | 25.5 | 16.5 |
| **8: Feb 22<sup>nd</sup>-28<sup>th</sup>** | 19.2 | 29.5 | 26.0 | 25.5 | 16.5 |

**Figure 2:**
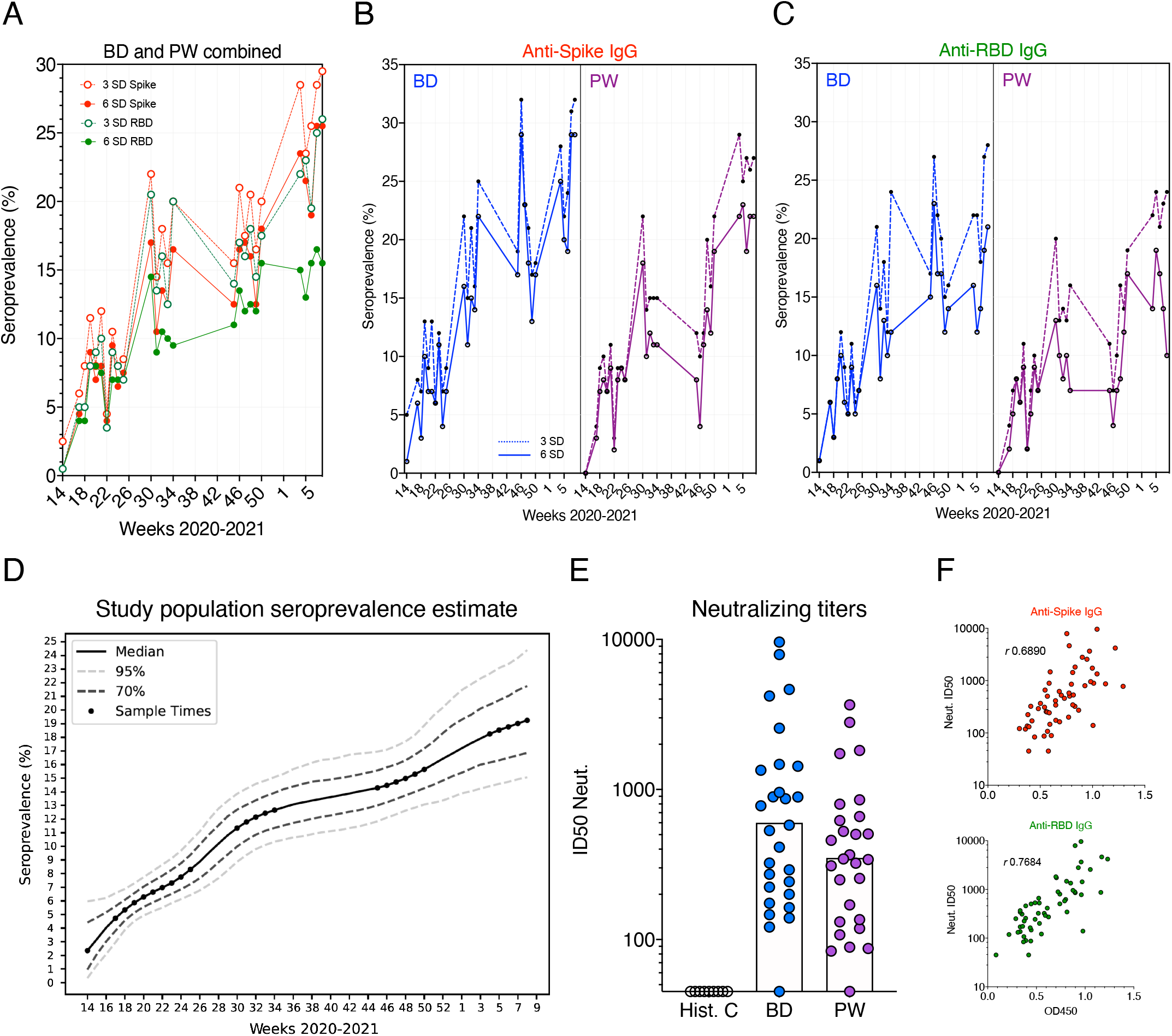
SARS-CoV-2 seropositivity estimates in Stockholm: March 2020-February 2021. **(A)** Sero-positivity estimates in BD and PW combined, according to 3 and 6 SD assay cut-offs. Simple linear regression was applied to each threshold. **(B)** Spike sero-positivity in BD and PW according to 3 and 6 SD assay cut-offs. **(C)** RBD sero-positivity in BD and PW according to 3 and 6 SD assay cut-offs. **(D)** Cut-off-independent Bayesian modelling of population sero-positivity. A frequentist, equal-weighted ensemble learner from the output of support vector machines and linear discriminant analysis was used to validate the Bayesian framework. **(E)** *In vitro* pseudotyped virus neutralizing titers in a subset of antibody-positive BD and PW. Bars represent the geometric mean. **(F)** Binding and neutralization -for samples in (E) -are highly correlated.

However, the many measurements between the 3 and 6 SD cut-offs for both or a single antigen (Fig. 1B and C) pose a problem when assigning *case* to low values; uncertainty that can significantly skew seroprevalence estimates and is undesirable at the individual level. Therefore, to provide an accurate seropositivity estimate for our population and to model changes over time, we developed a cut-off-independent, probabilistic Bayesian framework^9,31^ that models the log odds that a sample is antibody-positive based on anti-S and -RBD IgG responses in training data^31^; in this case SARS-CoV-2 PCR+ Covid-19 patients (*n*=136) across the clinical spectrum (including asymptomatic/mild cases)^9^. Anti-S and -RBD responses were present in all PCR-confirmed infections used as training data^9^.

Using this more quantitative approach that better considers the wide range of responsespresent in the population and shares information between sampling weeks, we found seropositivity to increase sharply at the start of the pandemic (Fig. 2D). By the time the Covid-19 death rate in the country was at very low levels during August, following the first wave peak, the seroprevalence trajectory increased at a steady, but slower rate in approach to the winter second wave (Fig. 2D) -in agreement with continued viral spread in the Stockholm population during a summer recess in cases and mortality, and consistent with persistent individual antibody responses over a 9-month period^18,32^.

By week 50 (13^th^ December 2020), our probabilistic approach identified 14.8% (95% Bayesian CI [12.2-18.0]) of the cohort to have been previously infected (Table 1 and Supp. Table 1). The seroprevalence level again increased more rapidly between mid-December and the end of February 2021, consistent with the winter peak in mortality and infections, and reached 19.2% (95% Bayesian CI [15.1-24.4]) of the population by last sampling. Thus, approximately one-in-five healthy adults in Stockholm showed evidence of past natural SARS-CoV-2 infection before the start of mass adult vaccinations in the country. An equal-weighted probabilistic SVM-LDA learner that we had previously optimized for ELISA measurements^9^ showed highly consistent results (Supp. Table 2).

Importantly, 96% of seropositive blood donors and pregnant women randomly sub-sampled (*n=*56 from March-May 2020 samples) had virus neutralizing responses in their sera (ID_50_=600; 95% CI [357 – 1,010] and ID_50_=350; 95% CI [228 -538], respectively, Fig. 2E-F), with titers comparable to those engendered by recently-approved Covid-19 mRNA vaccines that were shown to be protective in clinical phase 3 trials^33,34^. These observations support that asymptomatic/mild infection generates an antibody response that provides a first line of defense against potential re-exposures^35^, although inter-individual heterogeneity, environmental factors and different SARS-CoV-2 variants will have a role in individual outcomes^36–38^.

## Discussion

To characterize immunological responses to SARS-CoV-2 and safeguard public health, it is critical to monitor the level of population immunity after natural infection^5^, especially in settings with different public health measures – allowing for concurrent and retroactive evaluation of different strategies. As Sweden has taken a unique public health approach to mitigate the effects of the virus, data from the country provide an important contrast to comparable settings.

Serology is amenable to studies of large cohorts and remains the *gold standard* for determining previous exposure to pathogens. Several studies have highlighted the protective role B cells play in controlling SARS-CoV-2 infection in humans^35,39^ and animal models^40,41^, while potent neutralizing and convergent antibodies were rapidly isolated from infected donor samples^42–44^. Indeed, evidence suggests that most persons (>91%) previously infected (positive PCR test) with SARS-CoV-2 develop virus-specific antibodies, including following mild or asymptomatic infections^9,18,19,45^. Critical on-going and future research is required to determine the duration of B cell memory in those infected, as well as following vaccination.

Research shows that the majority of mildly infected individuals maintain a detectable virus-specific B cell response for at least 10 months^32,43^. Together with the steadily increasing seropositivity observed in this study and in the New York City (USA) population^46^, these findings help allay early concerns that immunity waned in the short-term; although individuals with mild infection can have antibody titers close to the assay boundary that decline from peak levels at different rates between individuals. Early results suggest that ∼10% of SARS-CoV-2 seropositive individuals can lose detectable titers 10 months post-PCR test^11,18,47^, although this proportion remains to be reported in larger and different cohorts. Therefore, the levels of past infection may be slightly higher than we report. Anti-SARS-CoV antibodies have been reported to persist for 3 years following more severe infection^48,49^, while detectable responses to seasonal coronaviruses generally wane within 1-2 years^50^. Importantly, as the activated B cell pool and antibody response contracts following viral clearance (and declines in function with age^51^), the identification of low but persistent antibody responses (that may be highly effective) remains a challenge using conventional serological methods.

Notably, we observed virus neutralizing titers in our cohorts to be comparable to those engendered by the first mRNA vaccines of Pfizer/BioNTech^33^ and Moderna^34^, supporting that natural infection generates neutralizing immunity to the infectious strain, as do studies of healthcare workers^39^ and elderly care home residents and staff^52^, in whom re-infection rates were substantially reduced. The potential endemicity and continued evolution of SARS-CoV-2 may alter the nature of protective immunity (e.g., possible re-infection with a different strain), and strain-specific antibody responses should be used to optimally coordinate vaccine interventions and inform public health measures.

Epidemiologically, the data presented here indicate that by the end of the first year since virus emergence in Sweden, approximately one-in-five blood donors and pregnant women in Stockholm had been previously infected by the virus.

This situation in Stockholm is similar to that observed in New York City, USA, where despite high rates of infection and mortality^53^, herd immunity was not attained within six months of the outbreak, with seropositivity reaching *∼*20% of the general adult population^46^. Despite an estimated 60% IgG seroprevalence after the first wave^54^, Manaus, Brazil was also not spared a severe second wave of infections^55^. As SARS-CoV-2 variants continue to emerge^56^, seroprevalence measurements reported from many countries highlight a continued need to curtail viral transmission through social restrictions and effective vaccination programs.

Our study supports that Stockholm has suffered a higher number of infections (following the first wave) than the majority of European locations studied^57–62^. However, direct comparisons between sites are complicated by differences in the demographics of the study subjects as well as the S&S of the assay used for antibody detection^63^. Longitudinal studies over longer timescales (first, second and future waves) and different cohorts are needed for an improved understanding of immune responses to SARS-CoV-2 infection.

## Conclusions

These data highlight a relatively high SARS-CoV-2 seroprevalence in Stockholm, Sweden following one year of community transmission. Population seropositivity informs disease epidemiology and public health approaches and can help target vaccines where they are most efficacious.

## Data Availability

Data generated as part of the study, along with custom code for statistical analyses, is openly available via our GitHub repositories: https://github.com/MurrellGroup/DiscriminativeSeroprevalence/ and https://github.com/chr1swallace/seroprevalence-paper.

## Author contributions

GKH and XCD designed the study, analyzed the data and wrote the manuscript with input from co-authors. JA, TA, SM and GB provided the study samples. LH, DJS, GM and BM generated SARS-CoV-2 antigens and pseudotyped viruses. XCD and JR generated the ELISA data. DJS performed the neutralization assay. MC, CW, N.F.G, and BM carried out statistical analyses. MC and BM developed the Bayesian framework.

## Acknowledgments

We would like to thank the study participants and attending clinical teams for making the research possible. Funding for this work was provided by a Distinguished Professor grant from the Swedish Research Council (agreement 2017-00968) and the National Institutes of Health, USA (agreement 400 SUM1A44462-02) awarded to GKH. CW received funding from the Wellcome Trust (WT107881) and Medical Research Council, UK (MC_UP_1302/5).

Additional funding was made possible by the European Union-funded CoroNAb project (coordination number 101003653). For the purpose of Open Access, the author has applied a CC BY public copyright licence to any Author Accepted Manuscript version arising from this submission.

---

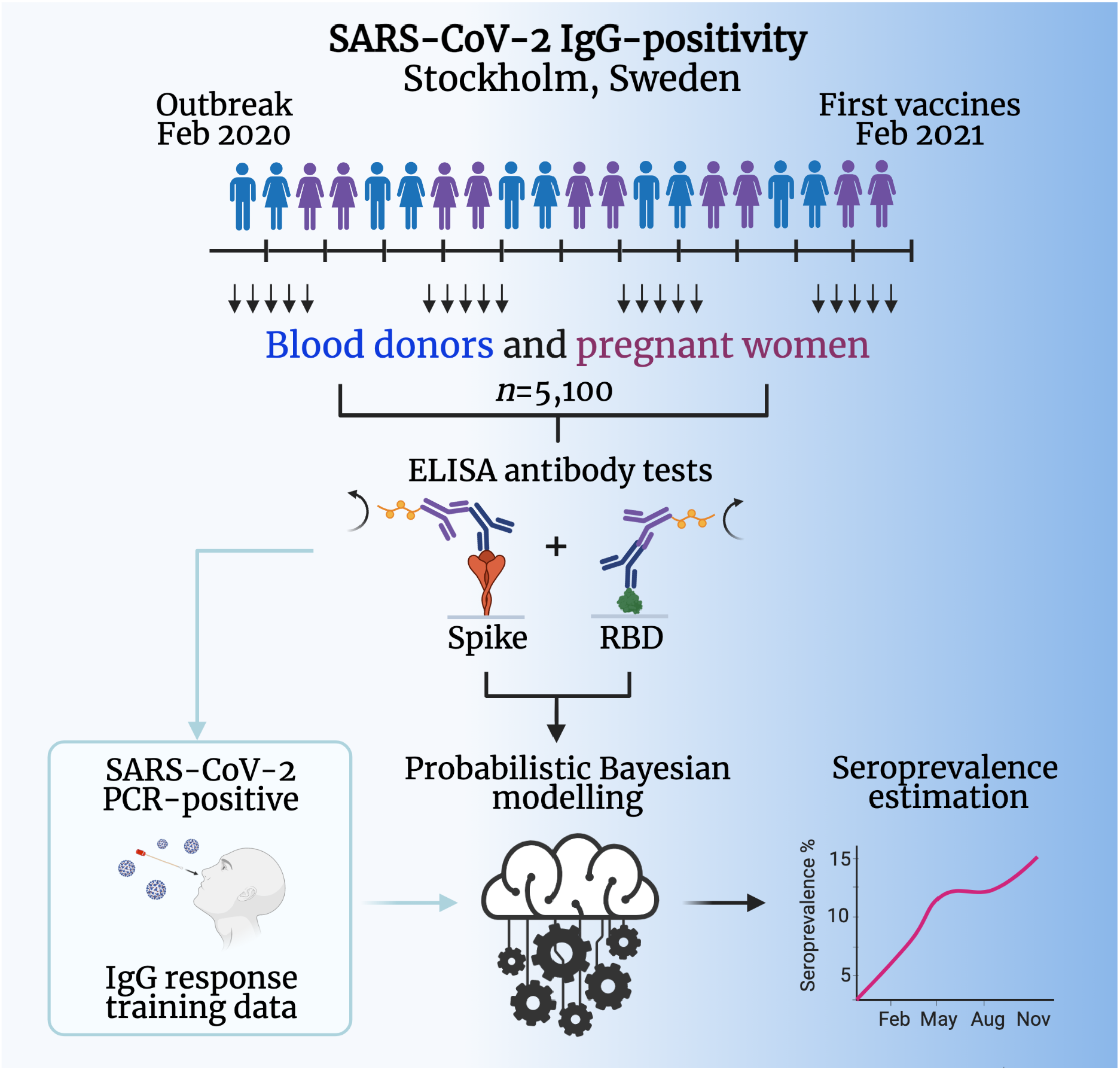

